# Excess mortality attributable to the 2025 Iberian Peninsula blackout

**DOI:** 10.1101/2025.06.03.25328877

**Authors:** Garyfallos Konstantinoudis, Julien Riou

## Abstract

On 28 April 2025, a widespread power outage affected mainland Portugal and Spain for around ten hours, causing major disruptions and eight reported deaths. Health impacts of blackouts range from direct effects, like fires and medical equipment failure, to indirect effects such as disrupted healthcare. We present the first systematic assessment of mortality linked to this event, disentangling impacts by age, sex, and region. Using advanced Bayesian models that accounted for temperature, holidays, and spatiotemporal trends, we estimated expected deaths during the blackout week. Comparing observed to expected mortality, we found limited excess deaths on the blackout day. In Spain, mortality rose over the next two days (+167 deaths; 95% Credible Intervals: +28 to +300), particularly among women aged 85+. No clear excess was found in Portugal. Regional differences suggest varying healthcare resilience. Findings highlight the underestimated health burden of blackouts and the need for improved preparedness during the climate crisis.

## Main

On 28 April 2025, electric power was interrupted across mainland Portugal and Spain for approximately ten hours, causing major societal disruptions and seven confirmed fatalities in Spain and one in Portugal [1, 2]. The event highlights the growing vulnerability of European power grids. Europe has previously experienced large-scale blackouts, including the Italy–Switzerland outage in 2003, the storm-induced Münsterland blackout in 2005, and the Adriatic coast outage in 2024 [3]. Such events are expected to become more frequent as European grids face mounting challenges in balancing supply and demand. These challenges stem from large-scale integration of renewables, declining fossil fuel capacity, increased exposure to cyberattacks, and the rising likelihood of extreme weather events—such as heatwaves, cold spells, floods, and storms—driven by climate change. Despite their growing frequency, the public health impacts of blackouts remain poorly quantified, particularly in terms of mortality risks associated with disrupted care, infrastructure failures, and cascading societal effects.

Power outages can affect health through a range of mechanisms. Injuries and fatalities may result from the use of candles, generators, or improvised heating devices indoors, as well as from carbon monoxide poisoning and fires [4, 5]. These pathways accounted for four of the eight officially reported deaths during the 2025 Iberian blackout [1], Disruptions to traffic systems—such as dark intersections or road lighting failures—can increase the risk of accidents [6]. Power failures can also disrupt healthcare services, delay emergency care, and pose serious risks to people who depend on electricity-powered medical devices [7]. Broader impacts, such as failures in water, sanitation, transport, and food systems, may further increase health risks, especially for vulnerable groups [8]. When blackouts coincide with extreme weather events, like heatwaves or cold spells, the health burden can escalate through heatstroke, hypothermia, and cardiovascular stress [9].

We present the first systematic assessment of the acute mortality toll of the 2025 Iberian blackout, with a focus on age, sex, and regional inequalities. We develop and validate 3 Bayesian spatiotemporal models and quantify the expected mortality across Portugal and Spain under the counterfactual of no blackout event. The models adjusted for key determinants of mortality, including temperature, the COVID-19 pandemic and public holidays, as well as seasonal and long-term temporal trends and regional dependences in daily death rates. By comparing observed all-cause mortality during and after the blackout to counterfactual expectations based on historical trends, we aim to capture the overall impact of the event, while disentangling individual and regional vulnerabilities.

We used mortality and population data from 21 contiguous regions across Portugal and Spain, covering a combined population of approximately 55 million. Bayesian models were trained on historical data from 2010 to 2024 to predict expected mortality for the first months of 2025, excluding the week of the blackout. We evaluated model performance and selected the specification that best reproduced daily mortality patterns by age group (*<*65, 65–84, and *>*84 years), sex (male, female), and region (NUTS2 level; see Online Supplement Figures S1–S2). Using the best-performing model, we then predicted expected mortality for the week of the blackout (28 April–4 May 2025), allowing us to quantify deviations from baseline.

We found limited evidence of significant excess all-cause mortality on the day of the blackout itself (lag0) in either country, with an estimated −32 excess deaths (95% CrI: −71 to +4) in Portugal and +14 excess deaths (95% CrI: −59 to +80) in Spain, Table 1. To account for potential delayed effects, we examined excess mortality over different, a priori selected, lag periods following the blackout. In Portugal, the results remained inconclusive: estimated excess deaths were +6 (95% CrI: −65 to +73) when considering deaths occurring within 0 to 2 days (lag0-2) after the event, and −19 (95% CrI: −154 to +94) when considering a longer lag window of in total 7 days (lag0-6). In contrast, in Spain, we found strong evidence of excess mortality within 2 days after the blackout, with an estimated +167 excess deaths (95% CrI: +28 to +300) compared to the counterfactual scenario without blackout. This represents a relative increase of +2.4% (95% CrI: +0.4% to +4.3%) (Online Supplement Table S1). However, when considering lag0-6, the evidence for excess deaths in Spain became inconclusive again.

**Table 1:** Estimates of excess all-cause deaths linked to the 2025 Iberian blackout of 28 April 2025 in Portugal and Spain by age group and sex, and with different time lags (0 days, only on the day of the blackout; 0-2 days, adding the next 2 days; and 0-6 days, adding the next 6 days).

|  |  | Portugal |  |  | Spain |  |  |
| --- | --- | --- | --- | --- | --- | --- | --- |
| Age | Sex | Lag 0 days | Lag 0-2 days | Lag 0-6 days | Lag 0 days | Lag 0-2 days | Lag 0-6 days |
| 65< | Males | +7 (-3, +17) | **+25 (+7, +42)** | +16 (-9, +41) | 0 (-21, +18) | +21 (-15, +53) | -10 (-64, +42) |
| 65< | Females | -3 (-11, +4) | -4 (-17, +7) | -9 (-29, +9) | +9 (-6, +22) | -1 (-27, +25) | -26 (-68, +12) |
| 65< | Total | +5 (-8, +16) | +21 (-1, +41) | +7 (-28, +39) | +8 (-17, +32) | +18 (-25, +59) | -36 (-103, +25) |
| 65-84 | Males | -5 (-23, +10) | 0 (-35, +31) | -7 (-66, +42) | -10 (-44, +23) | +7 (-65, +76) | -12 (-142, +124) |
| 65-84 | Females | -2 (-18, +11) | +3 (-25, +30) | -11 (-60, +36) | 0 (-28, +25) | 43 (-14, +96) | -6 (-113, +93) |
| 65-84 | Total | -8 (-32, +14) | +3 (-40, +43) | -17 (-92, +48) | -12 (-57, +33) | +50 (-38, +140) | -26 (-185, +142) |
| >84 | Males | -14 (-30, +1) | +3 (-26, +32) | 26 (-27, +75) | -4 (-32, +24) | +2 (-56, +60) | +5 (-115, +110) |
| >84 | Females | -14 (-34, +6) | -21 (-57, +17) | -34 (-107, +35) | +22 (-17, +54) | **+98 (+21, +167)** | +124 (-37, +265) |
| >84 | Total | **-28 (-54, -4)** | -17 (-62, +27) | -10 (-95, +77) | +17 (-30, +61) | **+99 (+6, +190)** | +127 (-70, +306) |
| Total | Males | -12 (-38, +12) | +28 (-15, +72) | +34 (-48, +109) | -15 (-64, +34) | +27 (-68, +127) | -20 (-204, +159) |
| Total | Females | -20 (-46, +5) | -23 (-70, +26) | -52 (-142, +35) | +29 (-21, +72) | **+139 (+39, +228)** | +92 (-108, +271) |
| Total | Total | -32 (-71, +4) | +6 (-65, +73) | -19 (-154, +94) | +14 (-59, +80) | **+167 (+28, +300)** | +72 (-202, +319) |

Figure 1 shows the range of counterfactual deaths (blue ribbon) and the factual mortality (black dots), across the different age, sex groups and countries. We observed weak evidence of reduced mortality among individuals aged 85 years and older on the day of the blackout in Portugal, with an estimated −28 deaths (95% CrI: −54 to −4), Table 1, followed by increased mortality in the next two days, Figure 1A. In aggregate, we found some evidence of increased mortality among people younger than 65 for lags 0 to 2 days (+21; 95% CrI: −1 to +41), primarily driven by males (+25; 95% CrI: +7 to +42). In Spain, there was limited evidence of excess mortality during the day of the blackout across all age and sex groups, but observed number of deaths exceeded the 95% credible interval of the expected counts on the following two days, Figure 1B. The evidence towards increased mortality was strong for lags of 0 to 2 days for people older than 85 years, with an estimate of +99 (95% CrI: +6 to +190), primarily driven by females (+98; 95% CrI: +21 to +167), Table 1. Over the entire week of the blackout (lags 0 to 6 days), we found no evidence of an increased mortality.

**Fig. 1:**
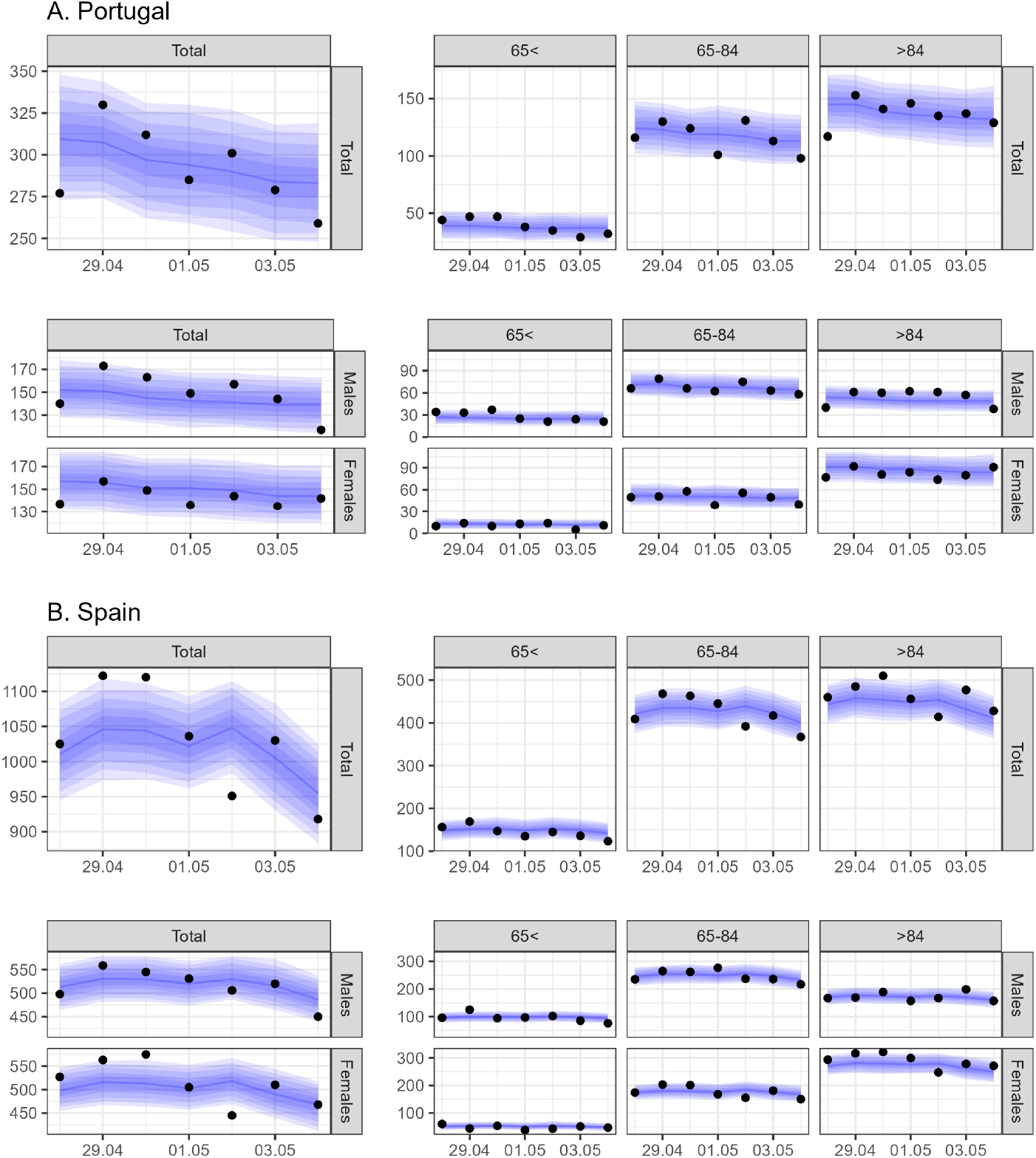
Expected daily all-cause deaths with 95% credible interval (blue ribbon) and corresponding observations of all-cause deaths (black dots) on the week of the 2025 Iberian blackout (28 April 2025 and the following six days), overall and stratified by age group and sex.

The geographical distribution of excess mortality across the regions of both countries revealed additional insights (Figure 2). The top panel of Figure 2 shows the percentage relative change in excess mortality, while the bottom panel displays the posterior probability that this excess is positive, reflecting the strength of evidence. In Portugal, the reduced mortality observed on the day of the blackout was mostly driven by the Algarve and Norte regions (north and south of Portugal in Figure 2A). However, accounting for lagged effects over the subsequent 2 or 6 days, evidence of increased mortality emerged in the central regions of Alentejo and Centro. In Spain, excess mortality on the day of the blackout was limited to the northwestern region of Galicia and the southern region of Andalusia (Figure 2). When considering lags of 0 to 2 days, the pattern became more geographically widespread (Figure 2). There are some indications that excess mortality even persisted up to 6 days after the blackout in some areas of the northern half of Spain (Figure 2).

**Fig. 2:**
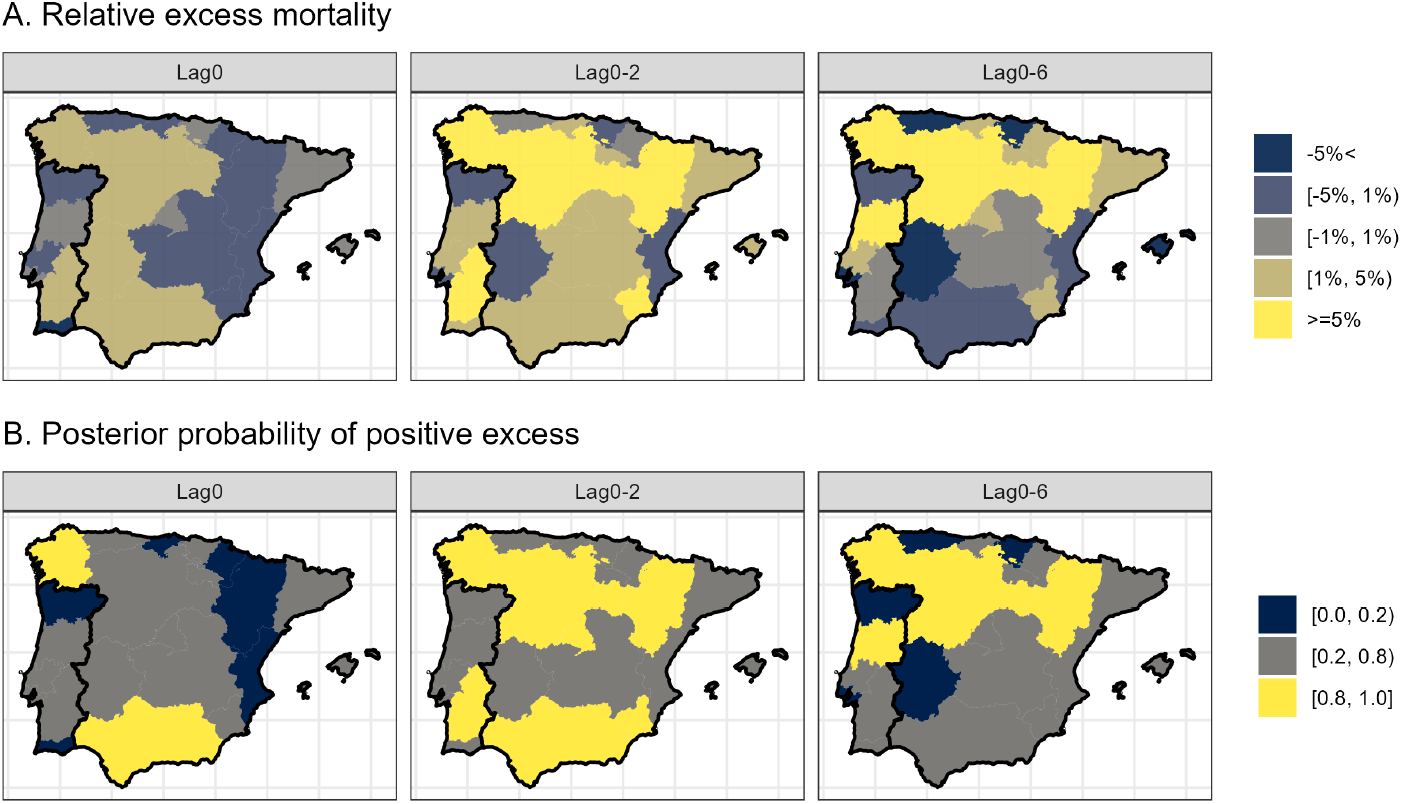
Relative excess all-cause mortality (panel A) and posterior probability of excess all-cause deaths greater than zero (panel B) by NUTS-2 region in Portugal and Spain with different time lags (0 days, only on the day of the blackout; 0-2 days, adding the next 2 days; and 0-6 days, adding the next 6 days).

Our results indicate that blackout-induced disruptions were associated with moderate increases in mortality, primarily observed or reported one to two days after the event. Based on these data alone, it remains unclear whether these excess deaths occurred on the day of the blackout and were identified with reporting delays, or whether they reflect lagged health consequences stemming from the disruptions. Nonetheless, our findings suggest that the total mortality burden—though moderate—substantially exceeds the seven officially confirmed deaths reported in Spain [1]. Given that such directly attributable fatalities are typically identified and reported promptly, it is plausible that the remaining 160 excess deaths reflect indirect impacts of the blackout, such as delays in emergency care, disruptions in chronic disease management, or environmental exposures.

The estimates of excess mortality varied across countries, regions or demographic groups, suggesting that the impact of the blackout was variable across contexts and settings. The concentration of excess mortality in females aged 85 or older in Spain points towards outpatient medical services, domiciliary nursing and care homes as particularly vulnerable settings. The absence of comparable effects in Portugal may reflect organizational differences. Indeed, Portugal has a highly centralized and integrated public long-term care system [10], whereas the Spanish system is characterized by decentralization and marked territorial disparities [11]. Additional research is needed to elucidate the differential impact of power outages on elderly populations dependent on electricity-powered healthcare infrastructures. We also found weak evidence of excess mortality among working-age males in Portugal, likely reflecting a modest rise in accidental deaths associated with the blackout. A few studies have documented short-term spikes in all-cause mortality during major power outages, on the day or in the subsequent few days [4, 12]. The 2003 Northeast U.S.A. blackout was linked to an increase of about 122% of accidental deaths and of 25% of non-accidental deaths, with older adults (65–74 years old) experiencing the largest mortality surge [6]. Other smaller power outages in New York City between 2002 and 2014 have also been associated with mortality increases of 2 to 6% in the following 1-3 days [13]. A 2019 typhoon-related blackout in Tokyo saw all-cause deaths increase by about 11% in the first three days post-event [12]. Such mortality impacts can be exacerbated when blackouts coincide with extreme weather events, which may explain the somewhat lower estimates reported for the 2025 Iberia blackout, which occurred in relatively mild weather conditions.

A key strength of our study is the rigorous methodological approach. The excess mortality framework proposed here is based on advanced Bayesian statistical modelling, which robustly captures direct and indirect mortality effects attributable to the blackout. This method has been widely used during the COVID-19 pandemic [14]. We also provide a thorough validation of the proposed model tailored to the specific context of daily all-cause mortality in Spain and Portugal in the spring of 2025, which underscores the high reliability of our model predictions in terms of bias and uncertainty. Finally, our results are based on updated population estimates, and account for the specific effects of the COVID-19 pandemic on all-cause mortality during the epidemic waves of 2020-2024. However, we face several limitations, including potential residual confounding due to unmeasured variables such as past or concurrent public health emergencies. This may lead to bias or increased uncertainty in the computation of expected deaths, which may in turn lead to limiting the sensitivity of our assessment of excess deaths. Likewise, the temporal and geographical granularity of our analytical framework may limit the detection of small-scale or localized effects. Last, out study lacks detailed data on the causes of death, which would have allowed insights into the sources of the observed variation in excess deaths.

In conclusion, our analysis underscores the significant health consequences of blackouts and power outages, particularly in the elderly, emphasizing the need for strengthened resilience and planning in healthcare infras-tructure. While the decarbonization of power systems is essential for climate mitigation, the April 2025 blackout reveals the fragility introduced by high renewable penetration without parallel investments in grid resilience. The health consequences of this event highlight the urgent need for policies that integrate renewable energy growth with robust system balancing, storage, and emergency health preparedness.

## Methods

We retrieved daily all-cause mortality data at the NUTS2 level for the period 2010–2025 from the National Statistics Institute in Portugal, and from the National Centre of Epidemiology at the Carlos III Health Institute, the Daily Monitoring Mortality System, the National Statistics Institute, and the Ministry of Justice in Spain. The number of deaths from all causes was available by sex and age. We grouped age into three categories: *<*65, 65–84, and *>*84 years.

Annual population figures for Portugal were obtained from the National Statistics Institute for the years 2011–2023, with estimates referring to December 31 of each year. For Spain, annual population data were obtained from the National Statistics Institute for the years 2010–2024, with figures consistently available for January 1 of each year. To estimate population counts for missing years, we applied a two-stage linear interpolation approach as previously described [14]. Briefly, we first used linear regression to estimate annual population figures for missing years (i.e., 2009, 2010, 2024, and 2025 for Portugal; and 2025 for Spain). We then performed linear interpolation to derive daily population estimates between 2010 and May 2025. Interpolation was conducted by age group, sex, and NUTS2 region. The resulting estimates were used as denominators in our analyses.

To support the prediction of daily counterfactual expected mortality, we included temperature and national holidays as covariates [15]. Temperature data were obtained from the ERA5 reanalysis dataset provided by the Copernicus Climate Data Store [16]. For each centroid of the 0.25°*×* 0.25° grid cells, we calculated the daily mean temperature for the period 2010–2025. These values were then aggregated to the NUTS2 level using population weights derived from the WorldPop dataset (www.worldpop.org), as they are available on a finer grid to match with the temperature data. We used the year 2018 for population weighting as the midpoint of our analysis. As mortality from all causes can vary during national holidays, we also included a binary variable indicating whether a given day is a public holiday, accounting for the regional holidays in Spain.

COVID-19 waves had a marked impact on mortality during parts of the 2020–2024 period. However, excluding the entire 2020–2024 period from the training set would introduce other challenges, particularly in capturing long-term mortality trends across age and sex groups. We therefore adopted a pragmatic approach, excluding only the weeks with substantial COVID-19 impact while retaining other weeks from this period to help anchor and characterize temporal mortality trends. Specifically, we used data on laboratory-confirmed COVID-19 deaths in Portugal and Spain from Our World In Data [17], and excluded from the training set any weeks with more than 100 such deaths in either country.

We considered three models to estimate the daily expected number of all-cause deaths, separately for each age and sex group and for the different countries (for simplicity we omit these indices in the following). Let *Y*_*slmt*_ denote the number of all-cause deaths and *P*_*slmt*_ the population in the *s*-th NUTS2 region on the *l*-th day of the week, *m*-th week of the year, and *t*-th year:

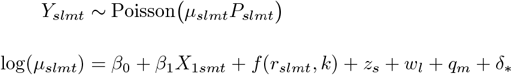

where *β*_0_ is the intercept, *β*_1_ represents the effect of national holidays, *f* (*r*_*slmt,k*_) captures the non-linear effect of temperature at lag k on all-cause mortality, *z*_*s*_ models spatial autocorrelation between NUTS2 regions, *w*_*l*_ captures day-of-week effects, *q*_*m*_ captures seasonality by week of the year, and *δ*_*\**_ represents the long-term temporal trend.

We considered the following specifications for *δ*_*\**_:

**Model 1**: *δ*_*\**_ = *u*_*mt*_, where *u*_*mt*_ is assigned an i.i.d. normal prior with no structure on the covariance matrix.

**Model 2**: *δ*_*\**_ = *u*_*mt*_ + *β*_2_*X*_2*mt*_, where *X*_2*mt*_ denotes the week and the year and *β*_2_ is a fixed linear trend.

**Model 3**: *δ*_*\**_ = *v*_*mt*_, where *v*_*mt*_ follows a second-order random walk process.

The non-linear effect *f* (*·*) of mean daily temperature *r*_*slmt*_ at lag *k* is modelled using distributed lag non-linear models [18]. The chosen basis functions for the space of temperature are quadratic B-splines with 5 degrees of freedom, with knots placed at 10°C, 18°C and 27°C. Similarly for the space of lags we used quadratic B-splines with 5 degrees of freedom, up to 21-day lags with knots placed at 5.25, 10.50, and 15.75. The choice of 21 is justified as the effects of cold temperatures are expected to be prolonged [18]. Spatial autocorrelation *z*_*s*_ is modelled with an i.i.d. normal prior without spatial structure, given the relatively large size of NUTS2 regions.

The models were considered in a Bayesian framework and fitted using Integrated Nested Laplace Approximation [19]. We assigned a normal prior with mean zero and zero precision to the intercept. For fixed effects, we used normal priors with mean zero and precision 0.001. For the hyperparameters governing the random effects’ precision, we applied penalised complexity (PC) priors [20]. Specifically, we calibrated the priors to ensure it is unlikely that relative risks exceed exp(2) due solely to spatial or temporal variation.

We trained the model using data from 2010 to 2025 (up to and including 27 April 2025) and generated predictions of area-level daily mortality for the week of the blackout, spanning from 28 April to 4 May 2025. To summarise the results, we retrieved samples from the posterior predictive distribution of daily expected mortality by age group, sex, and NUTS2 region for Spain and Portugal. We report the daily observed number of deaths during the blackout week alongside the corresponding posterior distribution of the expected number of deaths had the blackout not occurred. We also report the median and 95% Credible Intervals (CrI) of the cumulative number of excess deaths (observed minus expected) for three, a priori selected, time windows: (i) the day of the blackout (lag0), (ii) the blackout day plus two subsequent days (lag0-2), and (iii) the blackout day plus six subsequent days, to capture the entire week (lag0-6). Finally, we retrieved posterior estimates of the relative (percent) increase in mortality, i.e., the increase relative to the expected number of deaths had the blackout not occurred, by NUTS2 region.

To select the best-performing of the three considered model formulations, we performed a leave-the-last-week-out cross-validation. Briefly, we first defined the training set as the years 2010 to 2024, and the testing set as the first week of 2025. We then extended the training set to include the first week of 2025 and updated the testing set to the second week of 2025. This iterative procedure was repeated for both countries until all weeks up to and including 27 April 2025, one day before the blackout, were included. To compare model performance across the different cross-validation sets, we calculated three metrics: (i) bias, defined as the difference between the predicted and observed values; (ii) square root of mean squared error (MSE), calculated as square root of the average of squared differences between predicted and observed values; and (iii) coverage probability, defined as the proportion of observed values falling within the 95% credible interval of the predictions.

## Data availability

Population data in Spain can be retrieved from the Instituto Nacional de Estadistica in Spain (https://www.ine.es/jaxiT3/Tabla.htm?t=56945&L=0). Mortality data in Spain can be retrieved after request from the National Centre of Epidemiology at the Carlos III Health Institute. Mortality and population data in Portugal can be retrieved from the Instituto Nacional de Estadistica in Portugal (https://www.ine.pt/).

## Code and Reproducibility

All models were fitted using the Integrated Nested Laplace Approximation (INLA) using its R software interface [19]. Code for reproducing the analysis can be retrieved from GitHub (https://github.com/jriou/blackout-burden).

## Ethics

The study is about secondary, aggregate anonymised data so no additional ethical permission is required.

## Acknowledgements

G.K. is supported by an Imperial College Research Fellowship.

All authors acknowledge Infrastructure support for the Department of Epidemiology and Biostatistics provided by the NIHR Imperial Biomedical Research Centre (BRC). The authors also acknowledge Diana Gómez Barroso and Inmaculada León Gómez for preparing the mortality data in Spain.

## Author contributions

J.R. conceived the study. J.R. and G.K. developed the initial study protocol. G.K. developed the statistical model, prepared the population and covariate data and led the acquisition of mortality data. G.K. ran the statistical analysis for Portugal. J.R. and G.K. wrote the initial draft and all the authors contributed in modifying the paper and critically interpreting the results. All authors read and approved the final version for publication.

---

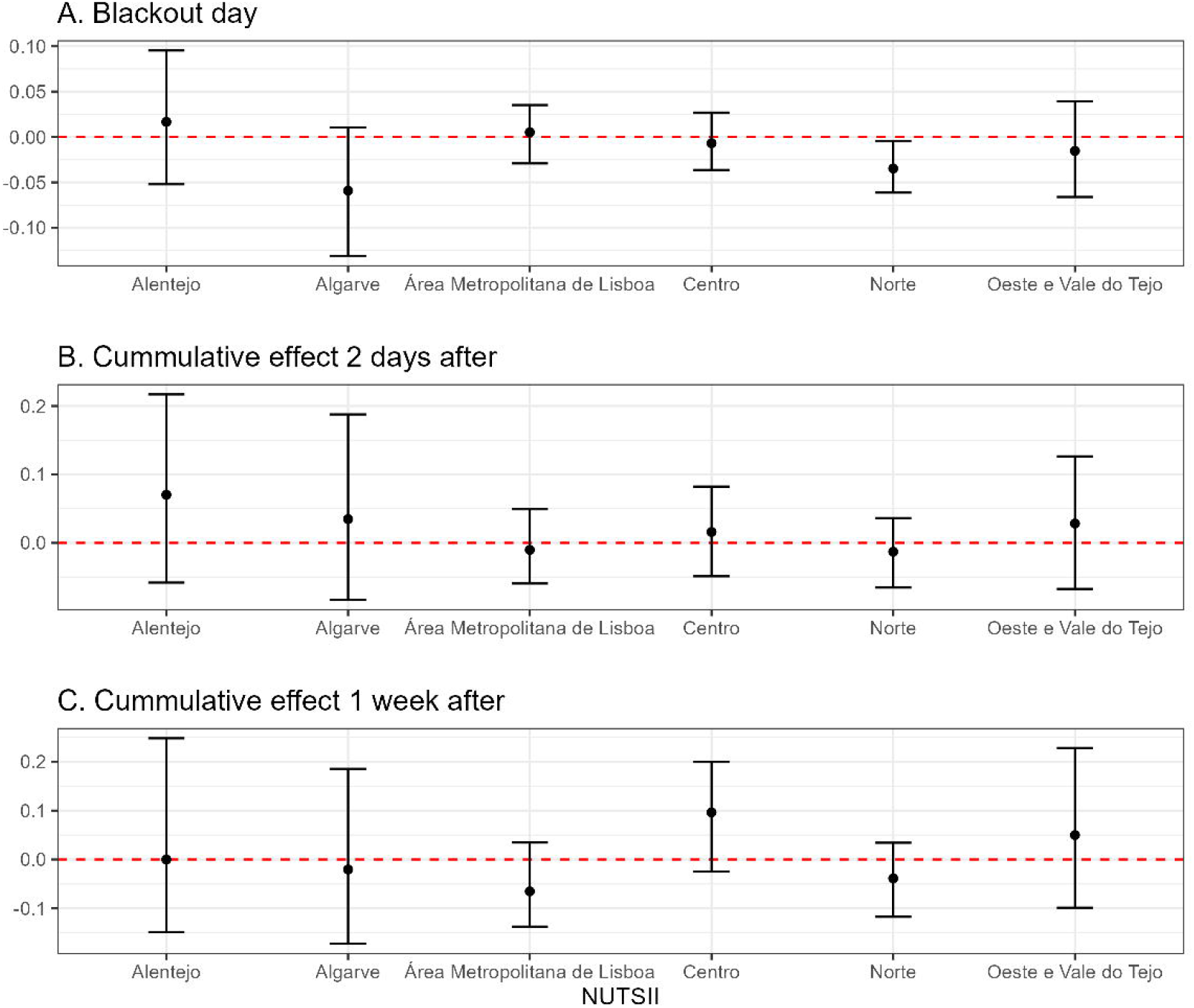

