## Supplementary material for "Excess mortality attributable to the 2025 Iberian Peninsula blackout": Online Supplement

Table 1: Estimates of percentage relative excess all-cause deaths linked to the 2025 Iberian blackout of 28 April 2025 in Portugal and Spain by age group and sex, and with different time lags (0 days, only on the day of the blackout; 0-2 days, adding the next 2 days; and 0-6 days, adding the next 6 days). The results are based on Model 3.

| Age | Sex | Lag0 | Pr | Lag0-2 | Pr | Lag0-6 | Pr |
|---|---|---|---|---|---|---|---|
| Portugal | | | | | | | |
| 65< | Males | 4.24% (-1.55%, 10.23%) | 0.91 | 14.20% (3.66%, 25.95%) | 1.00 | 8.94% (-4.41%, 26.62%) | 0.87 |
| 65< | Females | -3.14% (-11.32%, 4.71%) | 0.20 | -4.76% (-17.17%, 10.00%) | 0.23 | -10.47% (-27.36%, 13.24%) | 0.16 |
| 65< | Total | 1.89% (-2.98%, 6.62%) | 0.74 | 7.96% (-0.34%, 17.39%) | 0.96 | 2.64% (-9.33%, 16.74%) | 0.65 |
| 65-84 | Males | -1.11% (-4.70%, 2.23%) | 0.24 | 0.00% (-6.70%, 6.86%) | 0.47 | -1.47% (-12.34%, 9.84%) | 0.40 |
| 65-84 | Females | -0.64% (-4.96%, 3.33%) | 0.34 | 0.84% (-6.46%, 9.32%) | 0.58 | -3.10% (-14.86%, 11.69%) | 0.32 |
| 65-84 | Total | -1.01% (-3.70%, 1.72%) | 0.22 | 0.36% (-4.59%, 5.39%) | 0.54 | -2.05% (-10.17%, 6.27%) | 0.32 |
| >84 | Males | -3.83% (-7.92%, 0.27%) | 0.03 | 0.87% (-6.48%, 10.15%) | 0.58 | 7.37% (-6.65%, 24.68%) | 0.83 |
| >84 | Females | -2.38% (-5.39%, 0.99%) | 0.06 | -3.48% (-8.72%, 3.06%) | 0.14 | -5.55% (-15.60%, 6.43%) | 0.19 |
| >84 | Total | -2.86% (-5.40%, -0.40%) | 0.01 | -1.78% (-6.16%, 2.91%) | 0.23 | -1.03% (-9.03%, 8.74%) | 0.42 |
| Total | Males | -1.17% (-3.59%, 1.24%) | 0.16 | 2.78% (-1.48%, 7.73%) | 0.88 | 3.37% (-4.40%, 11.67%) | 0.80 |
| Total | Females | -1.90% (-4.21%, 0.50%) | 0.07 | -2.19% (-6.33%, 2.66%) | 0.18 | -4.94% (-12.43%, 3.63%) | 0.11 |
| Total | Total | -1.57% (-3.33%, 0.19%) | 0.04 | 0.29% (-2.97%, 3.69%) | 0.57 | -0.92% (-7.01%, 4.82%) | 0.38 |
| Spain | | | | | | | |
| 65< | Males | 0.00% (-3.01%, 2.70%) | 0.46 | 3.00% (-2.03%, 8.04%) | 0.87 | -1.46% (-8.66%, 6.64%) | 0.36 |
| 65< | Females | 2.49% (-1.68%, 6.34%) | 0.86 | -0.28% (-6.98%, 7.33%) | 0.45 | -7.18% (-16.84%, 3.70%) | 0.10 |
| 65< | Total | 0.78% (-1.63%, 3.05%) | 0.71 | 1.75% (-2.30%, 5.91%) | 0.80 | -3.39% (-9.25%, 2.54%) | 0.15 |
| 65-84 | Males | -0.59% (-2.46%, 1.39%) | 0.27 | 0.41% (-3.52%, 4.67%) | 0.58 | -0.69% (-7.59%, 7.73%) | 0.43 |
| 65-84 | Females | 0.00% (-2.19%, 2.11%) | 0.47 | 3.52% (-1.08%, 8.22%) | 0.92 | -0.48% (-8.40%, 8.17%) | 0.45 |
| 65-84 | Total | -0.39% (-1.88%, 1.12%) | 0.30 | 1.68% (-1.23%, 5.02%) | 0.86 | -0.87% (-5.88%, 5.04%) | 0.40 |
| >84 | Males | -0.32% (-2.58%, 2.10%) | 0.39 | 0.17% (-4.32%, 5.33%) | 0.52 | 0.42% (-8.71%, 10.05%) | 0.53 |
| >84 | Females | 1.14% (-0.88%, 2.96%) | 0.86 | 5.15% (1.03%, 9.33%) | 0.99 | 6.52% (-1.80%, 15.06%) | 0.93 |
| >84 | Total | 0.56% (-0.94%, 2.00%) | 0.75 | 3.20% (0.18%, 6.43%) | 0.98 | 4.09% (-2.12%, 10.47%) | 0.89 |
| Total | Males | -0.42% (-1.72%, 0.94%) | 0.28 | 0.73% (-1.80%, 3.68%) | 0.70 | -0.54% (-5.35%, 4.61%) | 0.42 |
| Total | Females | 0.83% (-0.58%, 2.10%) | 0.87 | 3.98% (1.06%, 6.78%) | 1.00 | 2.63% (-2.92%, 8.16%) | 0.81 |
| Total | Total | 0.21% (-0.81%, 1.13%) | 0.65 | 2.35% (0.38%, 4.31%) | 0.99 | 1.02% (-2.73%, 4.63%) | 0.70 |

Fig. 1: Cross validation results: Bias, coverage probability and square root of mean squared error across the different age and sex group for models 1, 2 and 3 in **Portugal**.

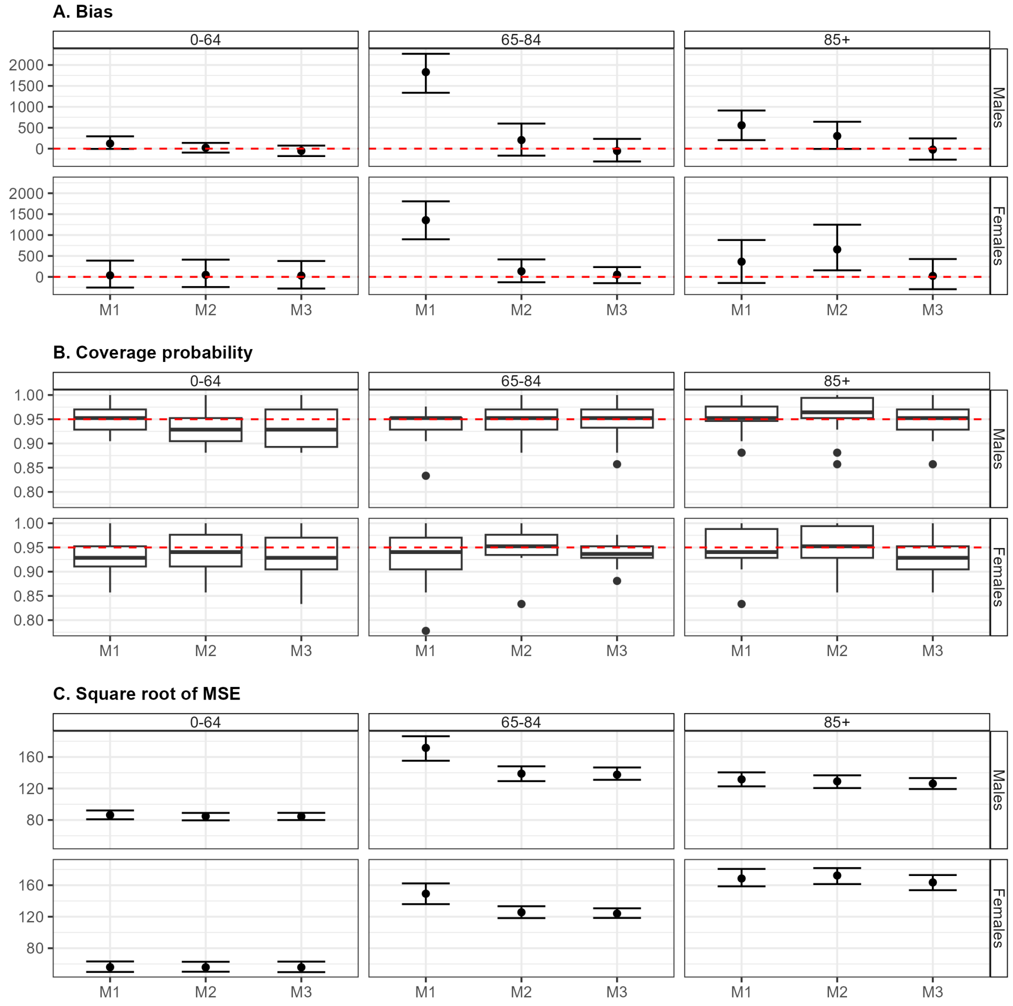

Fig. 2: Cross validation results: Bias, coverage probability and square root of mean squared error across the different age and sex group for models 1, 2 and 3 in **Spain**.

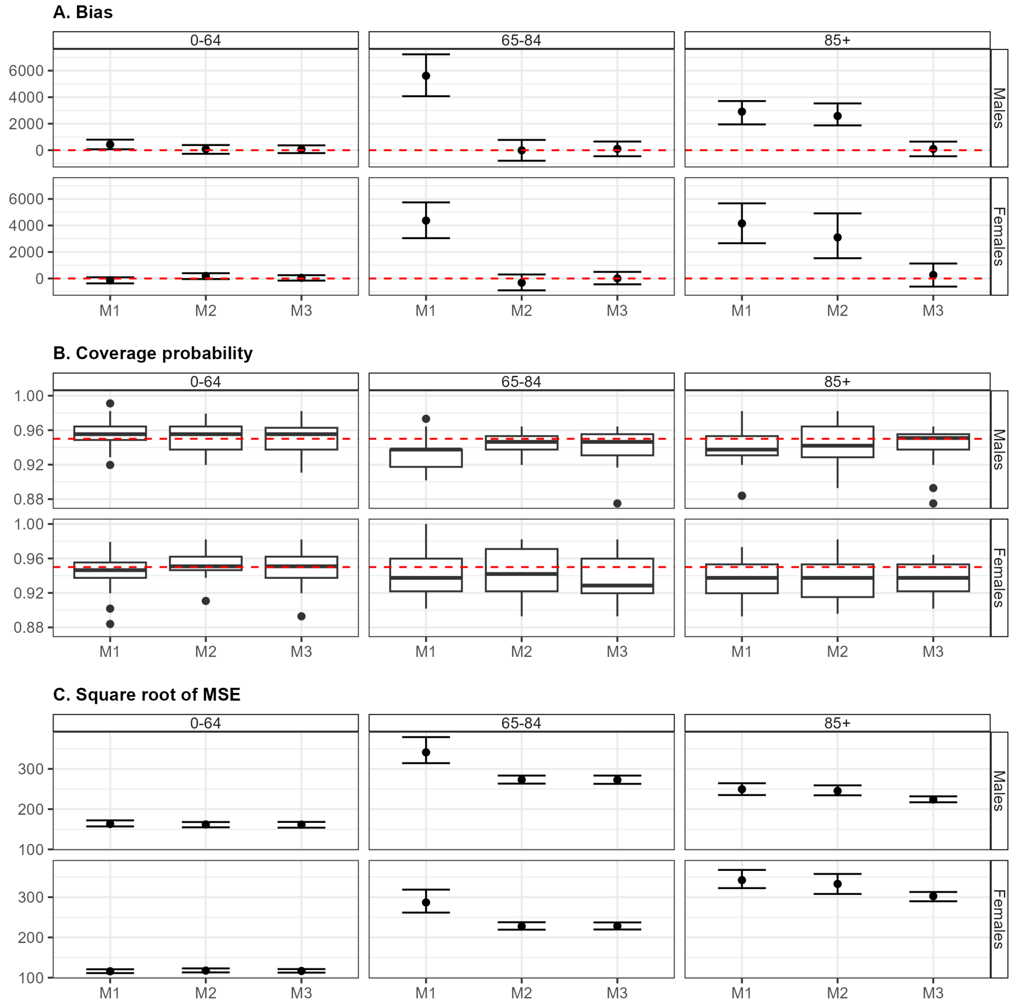
